# Genetic newborn screening stakeholder perspectives

**DOI:** 10.1101/2024.02.11.24302654

**Authors:** Didu Kariyawasam, Joanne Scarfe, Christian Meagher, Michelle A. Farrar, Kaustav Bhattacharya, Stacy M. Carter, Ainsley J. Newson, Margaret Otlowski, Jo Watson, Nicole Millis, Sarah Norris

## Abstract

**Background:** Newborn bloodspot screening is a well-established population health initiative that detects serious, childhood-onset, treatable conditions to improve health outcomes. With genomic technologies advancing rapidly, many countries are actively discussing the introduction of genomic assays into newborn screening programs. While adding genomic testing to Australia’s newborn screening program could improve outcomes for infants and families, it must be considered against potential harms, ethical, legal, equity and social implications, and economic and health system impacts. We must ask not only ‘*can’* we use genomics to screen newborns?’ but *‘should we’*?’ and ‘*how much should health systems invest in genomic newborn screening?*’.

**Methods:** This study will use qualitative methods to explore understanding, priorities, concerns and expectations of genomic newborn screening among parents/carers, health professionals/scientists, and health policy makers across Australia. In-depth, semi-structured interviews will be held with 30-40 parents/carers recruited via hospital and community settings, 15-20 health professionals/scientists, and 10-15 health policy makers. Data will be analysed using inductive content analysis. The Sydney Children’s Hospital Network Human Research Ethics Committee approved this study protocol [2023/ETH02371]. The Standards for Reporting Qualitative Research will guide study planning, conduct and reporting.

**Discussion:** Few studies have engaged a diverse range of stakeholders to explore the implications of genomics in newborn screening in a culturally and genetically diverse population, nor in a health system underpinned by universal health care. As the first study within a multi-part research program, findings will be used to generate new knowledge on the risks and benefits and importance of ethical, legal, social and equity implications of genomic newborn screening from the perspective of key stakeholders. As such it will be the foundation on which child and family centered criteria can be developed to inform health technology assessments and drive efficient and effective policy decision-making on the implementation of genomics in newborn screening.

## INTRODUCTION

Newborn bloodspot screening (NBS) is at an inflexion point in its evolution. A population health initiative, it has a historical mandate to identify newborns at risk of serious, childhood onset and usually treatable conditions, traditionally through analysis of biochemical indicators (1). The rapid advancement of genomic capabilities with the advent of next generation DNA sequencing methods has led to substantial interest amongst stakeholders to leverage these technologies within NBS (‘genomic NBS’). While the precise ways in which genomic NBS could be deployed are yet to be determined, a key artefact of any move to the use of genomics in NBS, is the potential to screen for many more genetic conditions than presently (2), with the goal of shortening an often arduous and complex diagnostic journey for affected individuals and their families (3). Avoiding a diagnostic odyssey is especially relevant for the diagnostically challenging 7000 or so rare non-communicable, genetic conditions (4), which collectively affect 8% of the Australian population, equivalent to 2 million individuals (5). For those affected with these conditions, there is typically a high level of symptom complexity conferring a significant health and psycho-social challenge, in part secondary to the accumulation of comorbidities associated with often delayed diagnosis. Identifying an increased risk of a genetic conditions through genomic NBS may lead to a timely diagnosis. A timely diagnosis, in turn, could enable expedient access to medical interventions and participation in research, facilitate early engagement with support, care and disability services and allow for financial and reproductive planning for affected families (6). Cumulatively, the effects of receiving an early diagnosis are manifold but essentially seek to improve health and psychosocial outcomes for the screened newborn (primary benefits) and as a downstream consequence, the wider family (secondary benefits) (1, 6).

These potential opportunities and benefits need to be balanced against possible harms to screened newborns and their families, as well as considered against the challenges that will arise from implementation at scale within complex health systems. If risks and barriers to successful use of genomic NBS are not identified and proactively addressed, the incorporation of genomics may lead to the erosion of an established and highly successful screening program (7). Moreover, it may risk existing high public trust in, and uptake of, NBS (8).

Proposed and implemented strategies for genomic NBS include identifying variations in single genes within first tier analysis (as deployed for NBS in spinal muscular atrophy) (9), using genomics as second tier screening for children with biochemical signatures of disease to reduce false positive rates (as proposed for NBS in Duchenne muscular dystrophy) (10), the use of targeted gene panels of varying sizes (11), or even whole exome or genome sequencing techniques for use as a health resource throughout one’s life (2, 12). For exome or genome sequencing techniques in particular, gaps in knowledge remain regarding their utility, cost effectiveness and feasibility for use at a population level.

Since 2022, the Medical Services Advisory Committee (MSAC), an Australian body that considers the addition of new conditions onto routine national NBS panels has evaluated and endorsed the use of first tier genetic technologies X-linked adrenoleukodystrophy, and continues to assess genomic NBS for a growing number of conditions ranging from hemoglobinopathies to lysosomal disorders (13). In line with the prevailing policy and international practice, MSAC continues to take what may be termed as a ‘*condition led*’ approach, as opposed to a ‘*technology led*’ approach to facilitate health technology assessments within newborn screening (14, 15). As such, conditions are individually evaluated for NBS endorsement through processes such as health technology assessments (HTAs). These processes focus on clinical and cost-effectiveness aspects of population screening, underpinned by evidence of test performance (rates of true/false positives and true/false negatives), acceptability of treatments for those identified as at risk through NBS, and an agreed policy on who, how and when to treat, often generated through pilot (NBS) programs that run over the course of many years (16). The creation of a robust evidence-base to inform these outcomes is time consuming, requires significant clinical research infrastructure and is especially challenging for rare diseases. Here, the development and application of HTA criteria is adversely impacted by small patient numbers within each condition, the lack of a coordinated strategy for data collection and synthesis, and less clinical evidence being available due to the challenges of conducting large scale clinical trials (17, 18). Furthermore, whilst HTAs recognise ethical, legal and equity aspects of genomics (including some early consideration of its role in population screening) these factors are currently only minimally integrated into the assessment processes (19). This is in contrast to growing support for incorporating such components early into health policy decision-making processes by stakeholders (20), reflecting an emerging paradigm of genomic NBS as a health intervention that is part of a ‘sociotechnical network’ of people *and* technologies, not separate from but requiring integration with clinical evidence of utility and incremental benefit from existing models of diagnosis and clinical care (21).

For future NBS programs to be flexible to accommodate the appropriate introduction of new health technologies such as genomics, decision-making must be efficient, robust and informed by all available evidence whilst also being contextualised for the specific challenges shared by many rare diseases (22), facilitated by the integration of real-world evidence, combined with expert input, clinical merit and public consultation (23). In tandem and in line with national policy, evaluating stakeholder perspectives of risk-benefit can help us understand the power and pitfalls of genomic capabilities in NBS, and inform a patient-driven and personalized model of care (24), (15). Synthesising stakeholder perspectives offers significant knowledge benefits including identification of conceivable benefits, risks, barriers and enablers of implementation. This evidence generation can help to streamline support and access to care by enabling co-design of an effective and holistic model of screening, diagnostic and clinical management. Concomitantly, this foundation may help to inform NBS policy and practice that is equitable, ethically appropriate and adapted to meet the needs of newborns and their families(15).

Whilst some of these issues have been explored in health systems with large private health insurance funding models, namely in North America (25–27), there is paucity of research on stakeholder perspectives of genomic NBS within health domains that are predominantly publicly funded, cover wide geographical expanses characteristic of the Australasian region, and encompass socio-demographically heterogenous (including Indigenous) populations. In Australia, while there has been a degree of systematic synthesis of information on genomic NBS from the perspective of the lay public (26, 28), and from rare disease medical experts (29, 30), the attitudes of parents of children with heterogenous genetic conditions (diagnosed with and without NBS) have not been analysed, nor have the unique challenges faced by policy makers when considering whether and how to incorporate genomics into NBS (31).

This study forms the first part of gEnomics4newborns research project: Integrating Ethics and Equity with Economics and Effectiveness for genomics in newborn screening (MRF2015965) (Figure 1). Accordingly, it will create knowledge on the perceived risks and benefits, barriers and facilitators of implementation of genomic NBS within an Australian context. This study will underpin the creation of a policy resource that will define key criteria for the assessment of new conditions onto routine NBS panels (screened through genetic technologies). Uniquely, this study will begin to inform policy decision makers on the priorities and relative importance of equity, ethical, legal, and social factors from a stakeholder perspective and how these can be incorporated into HTAs to maximise benefit, whilst mitigating harms of genomic technologies on a population level.

**Figure 1.**
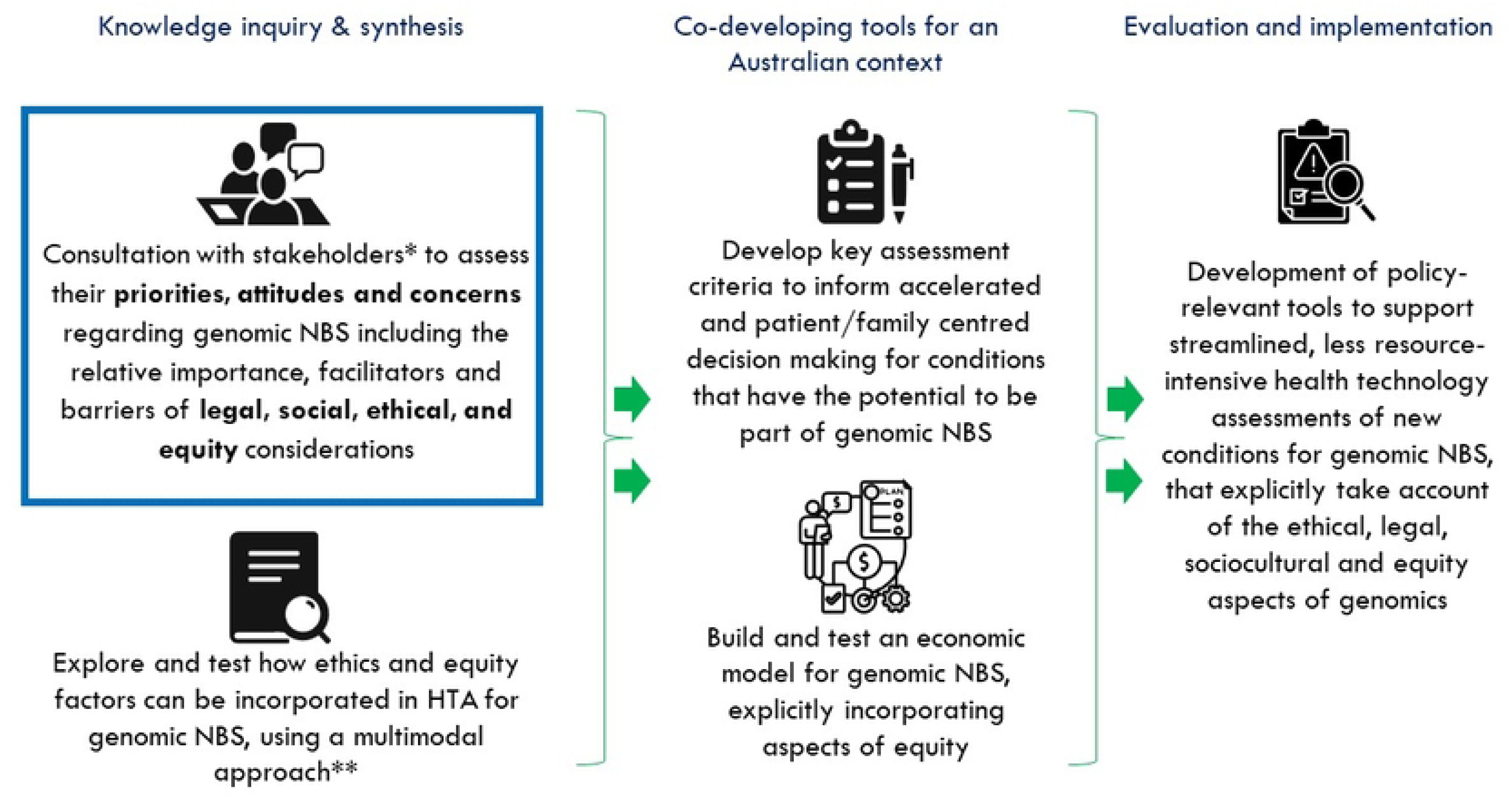
Placing the current study as the foundation of a research program to integrate ethical, legal, social and equity factors into health technology decision-making for genomic newborn screening. *Stakeholders include: Parents/carers, health care professionals/scientists, health policy makers. **Includes: Best-worst case scaling, Citizen’s Jury and Yarning Circle. Evidence generated through engagement with stakeholders will provide essential knowledge for a multimodal approach to generating recommendations. This will involve: 1. Best-worst case scaling, a process through which choice data from individuals is collated to understand choice processes using a top to bottom ranking method, 2. Citizen’s Jury, a deliberative inclusive approach to engaging with the community on complex issues. This process supports a group of randomly selected demographically diverse ‘jurors’ to hear and examine information given by expert witnesses on the complexities of genomics in NBS, and then to make recommendations about how policy makers should approach genomic NBS health technology assessment, 3. Yarning Circles, an established process of dialogue used within Aboriginal and Torres Strait Islander culture to assimilate and weigh information. This multimodal approach will lead to development of Assessment Criteria that can be used by future policy makers to establish methods of assessing new conditions in a faster, less resource-intensive manner and incorporating ethical, legal, social and equity aspects of genomic NBS.

## AIM

This study aims to explore stakeholders’ understanding, attitudes, concerns, and priorities related to genomic NBS in Australia. These stakeholders include parents and carers who have participated in NBS who have had one or more children diagnosed with genetic conditions, and those who have received false positive, false negative and uncertain newborn screen results; health care professionals and scientists; and health policy makers. This study will investigate a diverse range of perspectives on current and future impacts of the use of genomics in NBS, including the ethical, legal, social and equity implications, and explore how these views merge or diverge.

## METHODS

### Ethics approval

Ethics approval for the study is provided by the Sydney Children’s Hospital Network Human Research Ethics Committee [2023/ETH02371].

### Study design

This is a co-designed multi-cohort qualitative study using semi-structured interviews to explore understanding, experiences, and expectations of the potential use of genomics in NBS among parents/carers, health professionals and scientists, and health policy makers across Australia. Interview guides were developed following a scoping literature review of genomics in NBS, as well as input from the broader gEnomics4newborns stakeholders who have expertise in health and policy, clinical practice, consumer advisory and rare diseases advocacy groups. The full interview guides are included as Supplementary Material (S1 Appendix). In-depth, semi-structured individual and dyadic interviews will be held with parents and carers (n=30-40 participants), and individual interviews will be conducted with health professionals and scientists (n=15-20) and health policy makers (n=10-15). A thematic summary of the interview guides follows (Table 1). The Standards for Reporting Qualitative Research (SRQR) are being employed from study planning to conduct and reporting. (32).

**Table 1.** Thematic summary of interview guides.

| Domain | Discussion topics |
| --- | --- |
| *Parents / Carers – Cohort 1a (i) and 1b* |  |
| Experience of child's condition | Condition type and characteristics |
| Achieving a diagnosis | Experience of achieving a diagnosis through NBS |
|  | Experience of achieving a diagnosis outside of NBS |
| Care and management | Care and management of condition |
|  | Impact of condition on child and family |
| *Parents/Carers – Cohort 1a (ii)* |  |
| Experience of NBS | Experiences of participating in NBS |
|  | Experience of receiving a false positive or false negative NBS result |
|  | Perspectives on NBS following false positive or false negative result |
| *All parent/carer cohorts* |  |
| Genomic NBS | Understanding and interpretation of genomic NBS |
|  | Perceived risks of genomic NBS |
|  | Minimising harm to families through genomic NBS |
|  | Inclusion of conditions in genomic NBS |
| Model of care for genomic NBS | Consent to participate in genomic NBS |
|  | Delivery of results |
|  | Receiving uncertain results |
|  | Supporting families through achieving a screening result and diagnosis |
|  | Influence of genomics on acceptability of NBS as a public health program |
| *Health professionals / Scientists* |  |
| Professional experience | General |
|  | NBS and genetic diseases |
|  | Genomic sequencing |
| Genomic NBS | Perspectives on incorporating genomics into NBS |
|  | Included conditions |
|  | Perspectives on potential benefits and harms |
|  | Ethical, legal and equity considerations |
|  | Policy and implementation |
| *Policy makers* |  |
| Professional experience | Professional field |
| Genomic NBS | Views on the use of genomics in population screening and in NBS specifically |
|  | Perceived impact of genomics on current Australian NBS programs |
|  | Perceived benefits and challenges of genomic NBS |
| Genomic NBS policy | Impact on National Policy Framework |
|  | Views on current processes and required information when assessing genomic NBS |
|  | Ethical, legal, social and equity considerations |
| Genomic NBS implementation | Implementation challenges and perceived solutions |
|  | Impact of genomic NBS on parents/carers and families |

#### Sample and recruitment

Purposive sampling will be used to select participants and capture diversity in participant characteristics, including a range of (positive and negative) experiences across NBS and diagnosis. The number of participants will be determined through an iterative process of data collection, analysis and reflection, with data collected until theoretical saturation is reached and a range of perspectives are captured across participant types (33). Parents and carers of children diagnosed with a genetic condition, and parents and carers of children who received a false positive NBS result will be invited to participate in either individual or joint interviews with the other parent/carer of the child (defined as participant dyads). They will be recruited from hospital and community settings. All participants within this study are adults >18 years of age.

The study will recruit participants across a range of genders, sociodemographics and timing of screening and/or diagnosis. Non-English speakers will be supported by language interpreter services. Participants who identify as Aboriginal and/or Torres Strait Islander will be included in the study as they appear incidentally in the sample. These participants will be supported by the Aboriginal Hospital Liaison Officer and Aboriginal and Torres Strait Islander support services. Four cohorts will be recruited into the study:

##### Cohort 1a

Parents or carers who have participated in NBS in Australia and have: (i) children diagnosed within NBS programs with genetic conditions included in the current NBS Policy Framework (parents/carers of true screen positive individuals i.e. NBS screen positive and children go on to be diagnostically confirmed to have the condition) (17); or (ii) experienced false or uncertain results: a false positive result (NBS screen is positive but the child is NOT confirmed to have a genetic condition), a false negative result (NBS screen is negative for the named condition but the child develops symptoms of a condition and is later confirmed to have a genetic diagnosis) or a screen result with uncertain significance (i.e. those where there are variants of unknown clinical significance in disease causing genes).

#### Exclusion criteria

Parents/carers of children receiving a true screen negative result (parents/carers of individuals who are screened negative for conditions on the newborn screening panel and do not go on to be diagnosed with the condition at the time of the study) are excluded from the cohort, as the views of these individuals have been explored in previous research (34), and will also be captured in the other studies within the research program.

##### Cohort 1b

Parents or carers of children diagnosed with a genetic condition that would be unlikely to be (or has not been) approved under the current Australian NBS Policy Framework and associated decision-making pathways. Parents and carers who have been determined by their clinician to be at high risk of significant psychological distress by participating in the study will be excluded.

For these cohorts, clinicians from Sydney Children’s Hospital Network (SCHN) will pre-screen, select, contact, and obtain consent to be contacted by researchers from eligible participants from their clinical patient databases for parents/carers in Cohorts 1a and 1b. The managing clinician will be invited to put forward the name and contact details of participants who consent to be contacted to the study team. Potential participants will also be invited to enrol by study investigators when they are attending SCHN for routine clinical appointments as part of clinical care, where the managing clinician will obtain consent from prospective participants for the study team to contact them. To ensure diversity of opinions from across the community, patient advocacy and support organisations will also be approached to distribute advertisements for participation in the study via email and social media. Recruitment will be facilitated by partner consumer advocacy organisations in the rare disease community, the study’s Consumer Advisory Group, and project investigators. Parents/carers of children who received a false positive or false negative NBS result will be recruited via SCHN clinical databases. To avoid conflicts of interest, study investigators outside of the managing clinical team will recruit individuals into the study.

To ensure a diversity of opinions across the parent/carer cohorts, a recruitment screener will be used following the consent process to ensure that a diversity of participants and range of experiences are captured (S2 Appendix). A Decision Regret Scale will be administered during the screening process to capture the extent to which participants do or do not regret participating in newborn screening and, where relevant, consenting to their child/ren undergoing genetic testing (35).

##### Cohort 2

Health professionals (including midwives, genetic counsellors, and those experienced in NBS) and scientists involved in research and clinical care of children with genetic conditions and/or expertise in NBS and/or genomic medicine. Health professionals and scientists will be identified by purposive sampling relating to area of practice or expertise, experience, gender, age and ethnicity, with sampling aiming to represent stakeholders across Australia with at least one health professional/scientist from each state. Health professionals/scientists will be identified through investigator networks, contacted via email, and followed up with an additional email reminder or phone call two weeks later.

##### Cohort 3

Policy makers across Federal and State governments involved in NBS, genomics, and/or HTA. Policy maker recruitment will be facilitated by project investigators and researchers through existing networks.

For cohorts 2 and 3, passive snowball sampling may be used where interviewees will provide names of potentially suitable colleagues and information already in the public domain will be used to approach these individuals.

#### Data collection

Participants will take part in a single interview conducted by one researcher either face to face at a private location within hospital sites (such as an office or family room), or by videoconferencing as preferred by the participant. Parent/carer dyads will be interviewed together where possible. The managing clinician will not interview parents/carers of patients they are currently treating. Interviews will be audio-recorded with participants’ consent and will be 45-60 minutes in duration. If a participant experiences distress during or after an interview, a Psychological Distress Protocol will be implemented.

#### Data analysis

All recorded interviews will be transcribed verbatim and checked by the interviewer for errors. Transcripts will be managed using qualitative data analysis software NVivo V 14 and analysed using inductive content analysis (ICA). ICA is a method of analysing data where ‘content categories’ are generated from the data (36). ICA is appropriate for use where existing research is nascent, where an outcome of the research is to describe and understand the area being investigated, and where the research has direct importance for policy and practice (36). A proportion of early transcripts will be coded by two researchers independently. Preliminary coding will be compared on an agreed inductive coding frame that captures the breadth of views and experiences in the data. This coding frame will be applied to further transcripts independently by two researchers and revised iteratively as needed. Data from consumers, health professionals, scientists, and policy makers will be analysed separately, however where similar themes arise across the participant types this will be reported. Parent/carer dyads will be analysed as one unit (37). Reflexivity will be maintained by the research team through analysis and writing by recording, discussing, and challenging assumptions derived from their cultural, personal, and professional backgrounds (32).

## DISCUSSION

This will be a first-in kind study in Australia which focuses on the perspectives of genomic NBS from parents/carers whose children have been diagnosed with a genetic condition (both with and without NBS). Whilst previous studies have evaluated the perspectives of the public and professional stakeholders on genomic NBS in Australia (34, 38), the current study seeks to advance knowledge by assessing the perceived opportunities and challenges of using genomics in NBS amongst families with lived experiences of receiving a genetic diagnosis and evaluating how concordant or divergent their views are with other stakeholders including health care professionals, scientists and policy makers. This cumulative knowledge will be used to develop an integrated foundation on the key factors requiring consideration when making decisions on the clinical utility, validity and (unintended) ramifications of selecting conditions for genomic NBS.

As such, the study aligns with several pillars of the National Strategic Action Plan for Rare Diseases, which sets the impetus for collaborative research to facilitate effective clinical translation of health services (22). This study also reflects key national priorities that call for engagement of affected individuals and their families to help harness the current and future potential of genomics, facilitating processes for early diagnosis and ensuring that implementation of novel (genomic) technologies is equitable and fit for the needs of consumers (22).

Implementing genetic methodologies in newborn screening models undoubtedly provides a platform for the early identification of medically actionable conditions, with the prospect of changing disease trajectories and improving survival, function and quality of life for affected newborns (39, 40). As such, the drive to introduce novel genomic NBS programs is becoming increasingly apparent, with governmental initiatives, commercial and advocacy pressures all creating a wide and mounting array of conditions that are technically capable of being identified through population genetic screening in the newborn period (41).

However, underpinning these calls to action is a dearth of robust and high rigour evidence on the impact and potential risks and opportunities that such genomic NBS programs will create, particularly as genetic testing in and of itself has far reaching and long-term implications on not only affected individuals but also their families and the health system at large (42). Whilst many genomic tests are valued for the diagnosis and monitoring of conditions, their use on a population level (to screen for disease) has not always conferred a benefit on health outcomes, quality of life or survival (42). The limitations of a technology centric approach to NBS include ascertaining the predictiveness of genomic tests to account for the certainty and timing of disease onset for some conditions, (the latter leaving a population that requires proactive surveillance over the course of a lifetime, with the associated potential for psychosocial implications of ‘waiting for disease onset’) (43–45), the dangers of including *en masse* conditions in which there is no benefit from screening (46), the lack of reference genetic data for variants in ethnically diverse populations and the use of genetic screening for conditions where superior screening methods exist (42). These factors require careful evaluation, so that a barometer for the harms of screening can be in place when making health technology assessments, so as not to overstate the intended benefits.

Moreover, the processes that underpin genomics in NBS require specific consideration as issues of public health ethics. These include, but are not limited to, considering how a screening offer can be autonomously weighed by parents given the context of a likely publicly funded universal test offer, the influence of solidarity i.e. the concept of standing for, with and as newborns when thinking about the implications of this technology, rather than just being led by the existence of new (genomic) technology, and the collective values of reciprocity and trust to overcome inequities in access to novel technologies across the population (47). The potential for stigma and discrimination and legal imperatives to counter these effects are also important aspects to define and redress (48–50).

Through the evaluation and synthesis of stakeholder perspectives on the risks, benefits, attitudes and expectations from genomic NBS, the findings from this study will generate new knowledge to inform a model of genomic NBS that is adaptable to changing health technologies, effective and of high quality, incorporates processes to mitigate and address perceived harms and is placed within an ethically, legally and socially robust framework. Furthermore, this study will be used to develop the rationale and key criterion for implementing genomic NBS for conditions that differ in prevalence, disease characteristics and genotype across the heterogenous Australian population, to help overcome barriers to equity of access to novel (genetic) technologies and clinical care, as mandated by recent governmental initiatives and health policy directives (51). By using the perspectives of various key stakeholder communities to identify, consider and prioritise key utility, equity, ethical and social issues, the study findings will feed directly into methodological development in understanding the role of ethical, legal and social implications in HTA, improving transparency and fairness in health assessments (19, 52), whilst also placing stakeholder perspectives within the decision-making process, as a means to address perceived ethical and social complexities.

## Data Availability

No datasets were generated or analysed during the current study. All relevant data from this study will be made available upon study completion.

## ACKNOWLEDGEMENTS

This paper is submitted on behalf of the gEnomics4newborns Project Team including all chief investigators, associate investigators, co-researchers, and partners.

## AUTHOR CONTRIBUTIONS

The study was conceived and designed by SN, SK, JS, AJN, SC, MF and MO. SK, JS and CM wrote the first draft of the manuscript before receiving input from SN, AJN, SC, MO, MF, KB, JW, and NM. All authors have approved the final manuscript.

